# COVID-19 and depressive symptoms in students before and during lockdown

**DOI:** 10.1101/2020.04.27.20081695

**Authors:** Nicola Meda, Susanna Pardini, Irene Slongo, Luca Bodini, Paolo Rigobello, Francesco Visioli, Caterina Novara

## Abstract

The lockdown due to coronavirus pandemic may exacerbate depressive symptoms, experts argue. Here we report that students, a high-risk category for mental disorders, report on average worse depressive symptoms than six months before isolation. The prospective data reported herein should alert clinician of a possible aggravation as well as new-onsets of depressive symptoms in students.

---

The current coronavirus pandemic has been affecting Europe since late February 2020, forcing governments to put citizens in lockdown. Among growing concerns of the effects of isolation on mental health^1,2^, only retrospective data are available to assess if actual changes occur^3^. Here we provide prospective evidence of a change in depressive symptomatology of Italian students during COVID-19-related lockdown.

The study was approved by the University of Padova Ethical Committee of Psychology and participants provided informed consent. Between October 3^rd^ and October 23^rd^ 2019, we introduced the study to approximately 1000 University of Padova students, 153 of which matched target population characteristics (Italian native speaker students, age 18-30) and completed a demographic questionnaire and the Italian version of Beck Depression Inventory-2^4^ (BDI-2, a validated self- report questionnaire for depressive symptoms evaluation, the score of which correlates with severity of depressive symptomatology) online^5^, both in October and in April (between 3^rd^-23^rd^) 2020. We implemented generalised linear mixed models to evince if BDI-2 score changed during isolation with respect to the scores reported 6 months before. To assess a percentage change in BDI-2 score, we defined %ΔBDI-2 as the difference between BDI-2 score during lockdown and before lockdown, the whole divided by BDI-2 score before lockdown + 1 and analysed %ΔBDI-2 with linear mixed-effects models. To assess clinically relevant changes in depressive symptoms, we employed multinomial regression models. Sample characteristics and models employed are reported in Tables A and B, respectively. Anonymised dataset, further details on data analysis, and script are provided as Supplementary Material.

BDI-2 total score is slightly higher during lockdown than before (Figure, A and Table). We recorded that the median percentage increase is higher in males (+36%; IQR = −12 – 91%) than in females (+16%; −26 – 89%) and is independent from a history of mental disorder (Figure, B), although students with such history report higher before and during lockdown BDI-2 scores than students without any established diagnosis of psychopathology (Figure, C and Table). This increase is not significantly linked to sex, familiarity for a mental disorder, worry for one’s economic situation, or residence. Statistically, it is significantly linked to BDI-2 score before lockdown (Figure, D) and age, evidencing that younger participants with lower BDI-2 score before lockdown report higher percentage increases in BDI-2 score during lockdown. To assess if such increase could be clinically relevant, we divided participants into three clinically useful categories according to BDI-2 scores before lockdown (below 90^th^ percentile, above 95^th^ percentile, and between these two ranges^4^) and tested how many participants switched from one category to another, or remained in the same one during lockdown. We fit the observed data to a multinomial regression model and found that a median increase of 22% in BDI-2 score (IQR= −21 – 90%) would not clinically affect 79,2% of our target population (IQR = 74,7 – 81,4%); 8,2% (6,9 – 9,8%) would progress to a more serious clinical category (either from < 90^th^ to 90^th^-95^th^ range or from this latter to > 95^th^); and 6,2% (5,3 – 7,2%) would directly progress from < 90^th^ percentile category to the most severe clinical category (Figure, E and F). Less than 5% of participants would improve.

As Italy was entirely put in lockdown, it is impossible to assess isolation-independent changes in BDI-2 score. Students could be diversely affected by lockdowns: isolation may be responsible of a median increase of 22% in BDI-2 score, which would be clinically relevant for up to ≈ 15% of our target population. Our data should alert clinicians of possible aggravation of depressive symptoms in students, independently from a history of mental disorder.

## Data Availability

Dataset and R Script for analysis are provided as Supplementary Material

## Author contributions

All authors designed the study protocol, interpreted data and critically revised the manuscript; N.M. acquired data and analysed it and drafted the manuscript; P.R., F.V. C.N., S.P. provided technical, material or administrative support to the study; F.V., C.N., S.P. provided their supervision and expertise.

## Additional Information

Dataset and R Script for analysis are provided as Supplementary Material

---

**Figure Legend.**
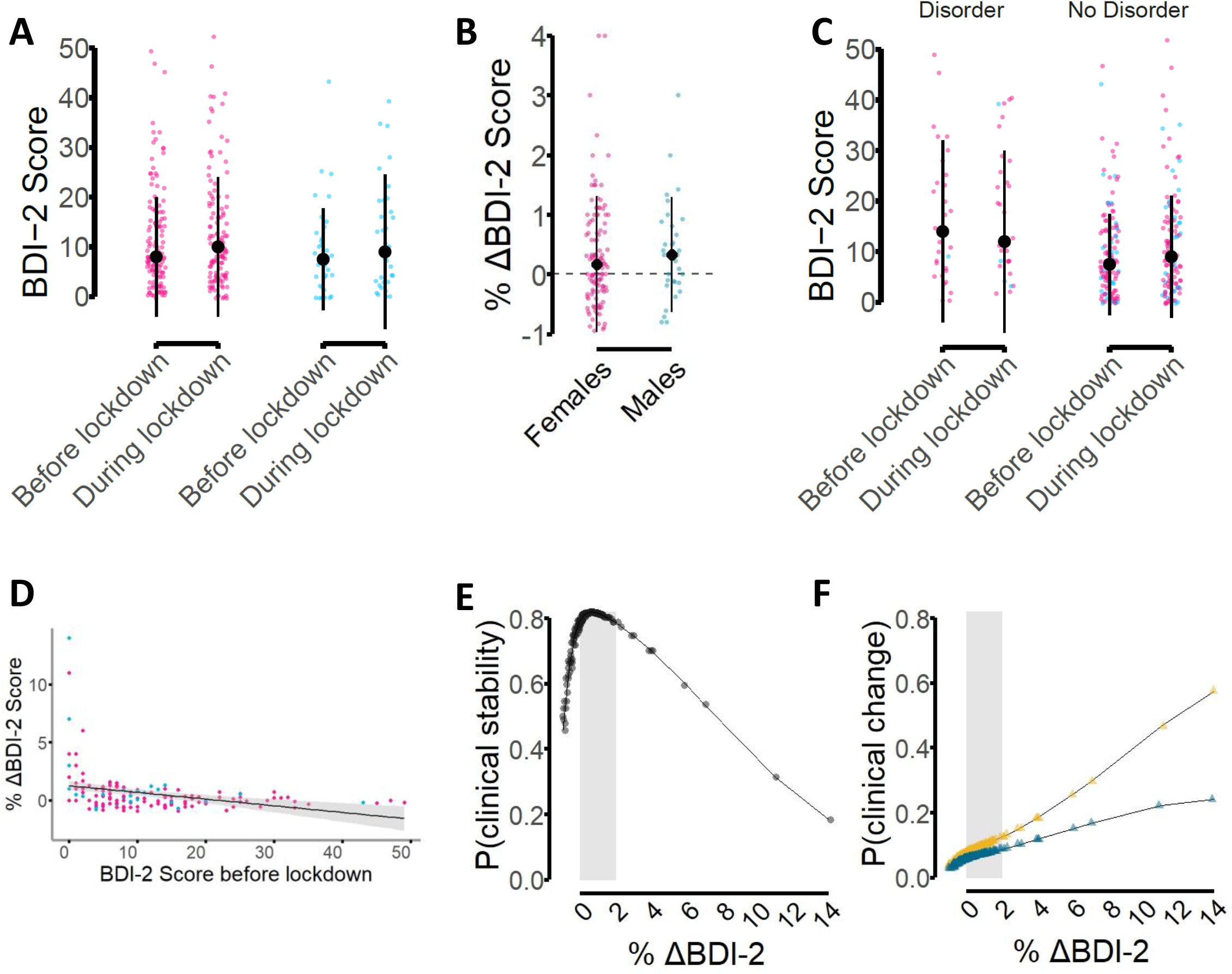
Possible COVID-19-related isolation impact on depressive symptomatology. Pink dots = females individual scores; blue dots = males individual scores. Pointrange represents median ± interquartile range. A, BDI-2 total score before and during lockdown. B, Percentage increase in BDI-2 scores. C, BDI-2 score stratified according to history of mental disorder. D, regression lines of percentage increase in BDI-2 score with respect to BDI-2 score before the lockdown. E, estimated probabilities of depressive symptoms stability (no clinical change) before- during lockdown as a function of percentage increase in BDI-2 score. F, estimated probabilities that depressive symptoms get worse (clinical category change); blue triangles = estimated probability of a steep worsening (from category below 90^th^ percentile, characterised by mild or no symptoms, to the most severe clinical category - higher than 95^th^ percentile); yellow triangles = estimated probability of worsening either from below 90^th^ percentile to 90^th^-95^th^ or from the latter range to above 95^th^ percentile; gray-shaded area = estimated probabilities for a 0-200% increase in BDI-2 score.

**Table.** Sample characteristics and regression models employed

| **Table. Sample characteristics and regression models employed** |  |  |  |  |  |  |  |
| --- | --- | --- | --- | --- | --- | --- | --- |
| **A Sample characteristics** |  |  |  |  |  |  |  |
|  | Females without disorder history | Females with disorder history | Females | Males without disorder history | Males with disorder history | Males | TOT |
| N (during lockdown) | 90 | 29 | 119 | 30 | 4 | 34 | 153 |
| Age (mean ± sd) | 22.0 ± 1.8 | 22.4 ± 1.5 | 22.1 ± 1.7 | 22.4 ± 2.3 | 22 ± 1.1 | 22.3 ± 2.2 | 22.2 ± 1.8 |
| BDI-2 score before lockdown (median; IQR) | 8; 4 – 15 | 14.5; 6.25 – 24.8 | 8; 5 – 17 | 7; 2 – 13 | 9; 9 – 9 | 7.5; 2.5 – 12.8 | 8; 4 – 16 |
| BDI-2 score during lockdown (median; IQR) | 9; 4 – 16 | 13; 8 – 26 | 10; 4 – 18 | 10.5; 3 – 18.5 | 6; 3.75 – 15.8 | 9; 3 – 18.5 | 10; 4 – 18 |
| % ΔBDI-2 (median; IQR) | 24%; -27 – 100 | 0%; -25 – 33 | 16%; -26 – 89 | 36%; -12 – 91 | 21%; -25 – 115 | 36%; -12 – 91 | 22%; -21 – 90 |
| **B Regression models** |  |  |  |  |  |  |  |
| **BDI-2 score model refers to Figure, A** |  |  | df | ΔAIC to null model | Significance of predictors |  |  |
| BDI-2 Score ~ Lockdown : Sex + Sex |  |  | 6 | 14.6 | Males:DuringLockdown, p < 0.001 (β = 0.27 ± 0.07)<br>Females:DuringLockdown, p = 0.019 (β = 0.08 ± 0.03)<br>Males, n.s. (p = 0.052) |  |  |
| **BDI-2 score model refers to Figure, C** |  |  | df | ΔAIC to null model | Significance of predictors |  |  |
| BDI-2 Score ~ DisorderHistory : Lockdown + DisorderHistory |  |  | 6 | 26.5 | No_DisorderHistory, p < 0.001 (β = -0.44 ± 0.12)<br>No_DisorderHistory:DuringLockdown, p < 0.001 (β = 0.19 ± 0.04)<br>DisorderHistory:DuringLockdown, n.s. (p = 0.18) |  |  |
| **%ΔBDI-2 model refers to Figure, B and D** |  |  | df | ΔAIC to null model | Significance of predictors |  |  |
| %ΔBDI-2 ~ BDI-2 score before Lockdown |  |  | 5 | 15.3 | BDI-2 score before Lockdown, p < 0.001 (β = -0.05 ± 0.01) |  |  |
| %ΔBDI-2 ~ Age |  |  | 4 | 2.26 | Age, p = 0.039 (β = -0.16 ± 0.07) |  |  |

Abbreviations and symbols: IQR = interquartile range; sd = standard deviation; null model =
regression model with no predictors; AIC = Akaike Information Criterion; ΔAIC = difference
between null model and better model AIC; df = degrees of freedom of a model; “:” means
interaction between predictors; β = estimated regression coefficient ± sd ;n.s. = not significant.
Further details on models can be found in supplementary material.

